# Role of Error Catastrophe in Transmission Ability of Virus

**DOI:** 10.1101/2022.06.28.22276997

**Authors:** Naoyuki Takahata, Hirotaka Sugawara

## Abstract

The role played by “error catastrophe” is explicitly taken into account in the mathematical formulation to analyze the COVID-19 data. The idea is to combine the mathematical genetics formalism of the error catastrophe of mutations in the virus gene loci with the standard model of epidemics which lacks the explicit incorporation of the mutation effect on the spreading of the viruses. We apply the formalism to the case of SARS-CoV-2 virus. We assume the “universality” of the error catastrophe in the process of analyzing the data. This means that some basic parameter to describe the error catastrophe is independent of which group (country or city) we deal with. Concretely, we analyze the omicron data of South Africa and then analyze the cases of Japan using the same value of the basic parameter derived in the South Africa analysis. The result of the excellent fittings of the two data, one from South Africa and the other from Japan with the common values of genetic parameters, justifies our universality assumption of these parameters.

## 1. INTRODUCTION

In usual analysis of virus spreading based on the standard model of epidemiology by Kermack and McKendrick (1927), we ignore the possible role played by mutations, at least in the explicit form. One of the present authors published two papers (Sugawara, 2021a, b) analyzing the world-wide COVID-19 situation based on the standard model through which he learned that the standard model does not incorporate the role played by mutations properly. Because of this fact, the analysis encountered some difficulty in interpreting the result of the analysis.

A simplified assumption of the absence of mutations in the virus genome is still made even for recently developed models in which, for example, multiple pathogen strains interact to each other through cross-immunity (Kucharski, Andreasen and Gog, 2016 for a review) or strains may interfere through the removal of individuals from the susceptible pool after an acute infection (e.g., Rohani et al., 2002). Meanwhile, the importance of random nature of pathogen mutations and emergence of new strains has been well recognized and studied by simulation under the assumption that a host’s risk of infection is proportional to the genetic distance between the infecting phenotype and the closest phenotype in the host’s immune history (Bedford et al., 2012). Recently, Miller, Elenberg and Dubrawski (2022) explored an existing agent-based model for influenza A/H3N2 which incorporates mutation effects into both transmission rate and the antigenic space in the standard model.

We present here a way to remedy the current situation for the presence of largely deleterious but occasionally advantageous antigenic mutations by introducing into the scheme of the standard model some elements showing the vital role played by mutations before, during and after the period of extensive virus spreading. When we analyze the data of variant strains of SARS-CoV-2 such as *α, β, γ, δ*, omicron BA.1 and omicron BA.2, we notice that the time variations of the number of the infected show certain interesting similarity among the different variant strains. In this regard, we emphasize an outstanding role played by error catastrophe (Eigen, 1971): Viruses can improve their transmission ability by mutations. But it seems that there is a maximum ability they can reach due to the errors during the DNA/RNA replication processes. After they reach the maximum (error threshold), the forced mutations (by such an enzyme as APOBEC3 (Sadeghpour et al., 2021) as was recently proposed) keep causing errors in the copying process of the virus multiplication and thus reducing the transmission power.

In this paper, we first explain the biological justification in more detail (section2). Then we formulate the process in mathematical terms (section 3). We apply the mathematical scheme developed in section 3 to actual data analysis taking the Tokyo case as an example (section 4). Section 5 is devoted to the analysis of omicron data from South Africa and Japan. The appendix deals with the issue of symbiosis in our formalism.

## 2. BIOLOGICAL JUSTIFICATION OF THE MODEL

As stated in the introduction, the analyses of the data of variant strains of SARS-CoV-2 show that the time variations of the number of the infected imply certain interesting similarity among the different variant strains. So far no clear explanation of this phenomenon exists within the framework of mathematical epidemiology. The data also show rather similar decaying patterns of the infection numbers of a certain variant once they reach the maximum in various countries where different epidemic policies are adopted including the vaccination rates. These similarities especially of the decaying patterns which are independent of the kind of variant strains or of epidemic policies seem to be explainable by assuming that the reason is on the side of viruses concerned.

A possible model is what we call an error catastrophe model (Eigen, 1971) which claims that the accumulated “error” in the mutation makes it hard for the multiplication process to be smoothly performed. The infection rate of such a virus as SARS-CoV-2 which has a rather high rate of mutations will be strongly influenced by the errors and its decaying patterns are largely determined by them. In the case of SARS-CoV-2, which allows APOBEC3 enzymes to directionally mutate from C to U, the error catastrophe may be particularly important. We do not need to treat this RNA editing by APOBEC3 distinctly from the usual mutation that occurs during the replication (Sadeghpour et al., 2021). It simply emphasizes the importance of “error catastrophe” and, therefore, it is hidden in our mathematical formulation.

The genome size of SARS-CoV-2 is about 30 kb, three times larger than that of typical RNA viruses such as influenza viruses and HIV. Despite being an RNA virus, CoV-2 accumulates mutations comparatively slowly under high fidelity of replication by expressing a 3’-to-5’ exoribonuclease nsp14-ExoN (Smith et al., 2013). However, if nsp14-ExoN is disrupted by self-mutagenesis or external agency, the mutation rate may increase 15 times (Robson et al., 2020; Thakur et al., 2022 for a review). It is observed that nearly one-half of nucleotide changes in CoV-2 are C to U transitions and occur preferentially in both 5’U/A and 3’U/A flanking sequence contexts that are favored motifs of the human APOBEC3 protein family of cytidine deaminases. As a result, CoV-2 may mutate at a rate of 3 × 10^−4^ /site/year, or 9/genome/year, consistent with a mean of 5.5 to 9.5 nucleotide differences between variants (Simmonds and Rampant, 2020; Ratcliff and Simmonds, 2021). Such high and context-dependent mutations likely hinder proper self-replication, lower the power of infection, though by yet unknown molecular mechanisms, and allow maladaptive amino acid changes to accumulate, leading to suicidal explosion of the viruses or error catastrophe. Even so, antigenic escape can occur and challenges pandemic control efforts. This unique paradigm of CoV-2 genome evolution is not incorporated by standard models of molecular epidemiology.

In the past, some efforts have been done to incorporate the pathogen evolution to the framework of epidemic models (SIR or its extension). For example, see section3 of Kucharski, Andreasen and Gog (2016). Compared to those works quoted there, our work is unique in its emphasis on the error catastrophe.

## 3. MATHEMATICAL FORMULATION

The purpose of this section, in a way, is to combine mathematical formulation of the mutation process with the standard model of mathematical epidemiology (Kermack and McKendrick, 1927; Kucharski, Andreasen and Gog, 2016). We start with a formulation of the mutation process and properly combine it with the standard model of epidemiology. Our target space is a well-defined population of humans such as a country or a city. We concentrate on the genes which are relevant for the transmission of the virus. This means that we concentrate mostly on the mutations in the spike proteins but not necessarily restricted to them.

We start by defining *x*_*i*_ to be the number of viruses that belong to the *i*-th variant. Here the “variant” means not just a prominent variant such as *δ* or omicron, but any group of viruses which has distinctive mutations from the original one. Let *a*_*i*_ denote the rate at which variant *i* reproduces (*a*_*i*_ does not necessarily mean the infection rate of variant *i*), and let *Q*_*ij*_ denote the probability of a virus of variant *i* mutating to variant *j*. Then the rate of change of *x*_*j*_ is given by

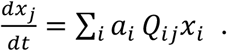

Suppose we group one kind of variants which have larger infection rates into type *X* and those variants which have smaller infection rate into type *Y*. We respectively denote *y*_*i*_, *b*_*i*_ and *R*_*ij*_ rather than *x*_*i*_, *a*_*i*_ and *Q*_*ij*_ for the latter case.

Then, we obtain

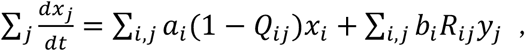

and

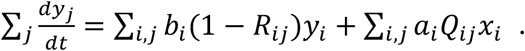

To simplify the discussion, we assume *a*_*i*_, *b*_*i*_, *Q*_*ij*_, and *R*_*ij*_ are all independent of suffix “*i*”. Then we get

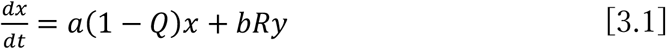

and

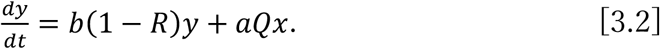

Here we have

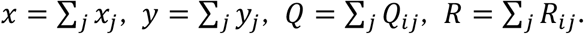

In the two-component vector form, Eqs. [3.1] and [3.2] become

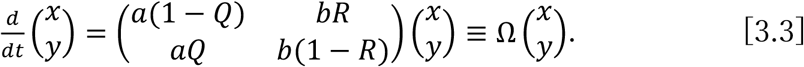

Here, 1+*a* is the average reproduction rate of variant in type *X* and 1+*b* is the same for type *Y*. Both *a* and *b* can be positive or negative considering the replication error. We note that all the parameters in the matrix Ω can depend on time *t* in principle. We however treat them as if time independent in this section. A slow time dependence will be introduced in the eigenvalues of matrix Ω in the actual analysis of the data as a kind of adiabatic approximation.

The number of the infected *I* at time *t* of our target population will change according to the following formula:

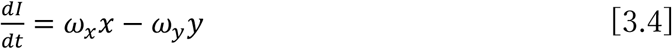

where *ω*_*x*_ and *ω*_*y*_ are constants. The eigenvalues of matrix Ω are given by,

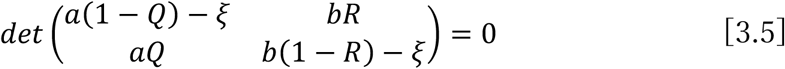

which gives

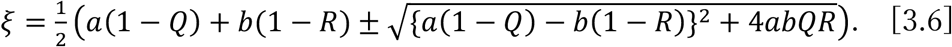

In general, *ξ* is a complex number:

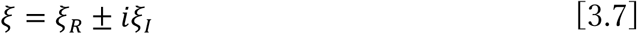

with

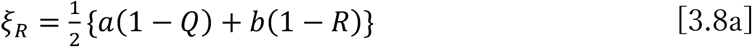

and

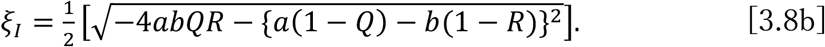

We get

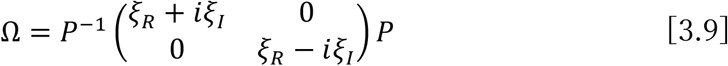

where *P* is the rows of the eigenvectors of Ω. Eq. [3.3] gives

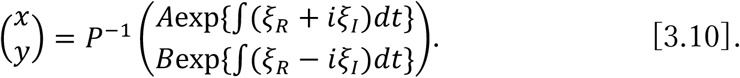

The general real solution is given as

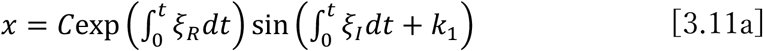

and

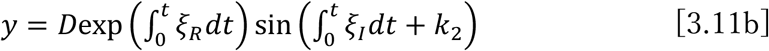

where *C* and *D* are constants. We may assume that the time dependence of *ξ*_*R*_ + *iξ*_*I*_ is not large and we ignore the contributions after the linear term in the power series expansion of *ξ* in terms of *t* for the later analysis of the data. Although a more precise treatment such as the perturbation theory developed in particle physics is available, it will be explored elsewhere. We note that we have

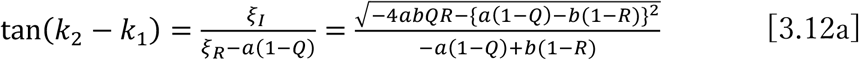

and

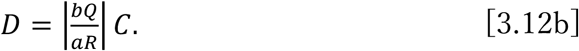

### Principle of error catastrophe

We see from Eqs. [3.11] that both *x* and *y* have an exponential factor 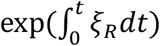. This means (due to Eq. [3.4]) that, for positive *ξ*_*R*_, the infection number behaves wildly, oscillation becoming larger and larger in its size. For vanishing *ξ*_*R*_, we have a periodical oscillation just like in regular influenza. The error catastrophe is defined to be the case when we have *ξ*_*R*_ < 0. A condition of this to happen for time-independent *ξ*_*R*_ is given by

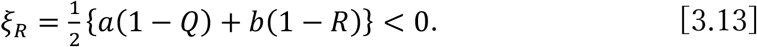

We assume that Eq. [3.13] is valid for SARS-CoV-2 virus at least for *x* > *x*_0_ (error threshold). There will be periods where this condition is not valid especially at the beginning of a surging of certain variant virus. The change of the infection number given in Eq. [3.4] is written as

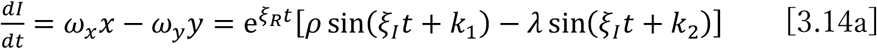

and

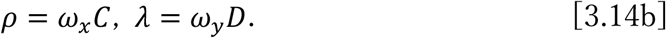

Here, for simplicity we assumed the eigenvalues to be time independent. Just to see the behavior of *dI*/*dt* for a positive or a negative *ξ*_*R*_, we plot Eq. [3.14] in Fig. 1a and 1b for two cases:

**Figure 1.**
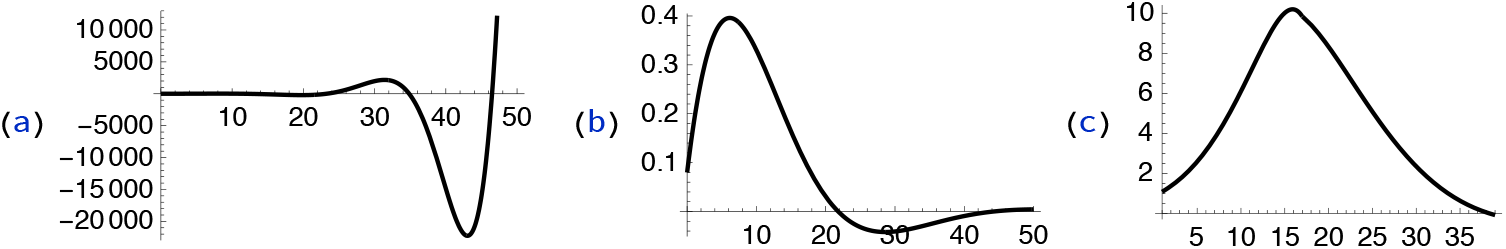
(a) Behavior of Eq. [3.14] for *ξ*_*R*_ = 0.2, *ξ*_*I*_ = 0.1, *ρ* = 5, *λ* = −0.1, *k*_1_ = 0, *k*_2_ = 0.83. (b) Behavior of Eq. [3.14] for *ξ*_*R*_ = −0.1, *ξ*_*I*_ = 0.1, *ρ* = 1, *λ* = 0.1, *k*_1_ = 1.0, *k*_2_ = 0. (c) Eq. [3.14] with *ξ*_*R*_ = 0.2 for 0 ≤ *t* ≤ 17 and *ξ*_*R*_ = 0.2 for 17 ≤ *t* ≤ 39. The other parameters are common: *ξ*_*I*_ = 0.1, *ρ* = 1, *λ* = 0.1, *k*_1_ = 1.0, *k*_2_ = 0.

These values correspond to the following basic parameters:

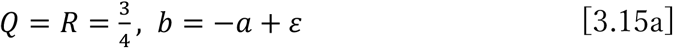

with

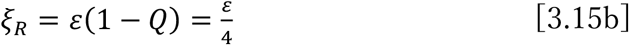

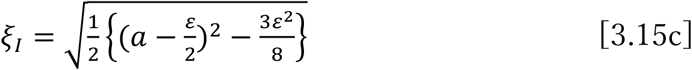

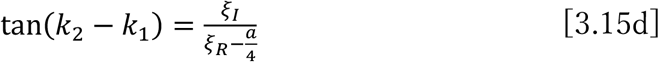

and *ε* = 0.8 for the case (a) and *ε* = −0.4 for the case (b). We also plot Eq. [3.14a] when *ξ*_*R*_ changes from 0.2 to −0.1 in the middle of peaking of the spreading. Concretely, we choose (a) initially, and (b) right after the peaking of the spreading. We see from these plots that:

1. There is a maximum value for the change of infection number for either case of *ξ*_*R*_ > 0 or *ξ*_*R*_ < 0.
2. Whether the positive *ξ*_*R*_ is kept (a) or it changes to a negative value (b), that is, whether *dI*/*dt* chooses the case depicted in Fig. 1c depends on the actual data.

Incidentally, we observe that any country or city has experienced a plural number of peaks in the COVID-19 daily infection number maybe except for China. It shows that each peak is caused by a new variant which is sometimes drastically new, such as *δ* type or omicron type, and sometimes just a small number of mutations different from the preceding variant.

Incorporating the standard model of epidemiology

Our method to combine the above formalism on mutation and the standard model of epidemiology is straight forward as shown here. The standard model (SIR) (Kermack and McKendrick, 1927) is defined to be the following set of differential equations for *S* (susceptible), *I* (infected), and *R* (recovered):

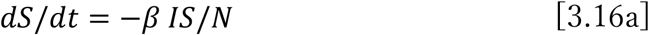

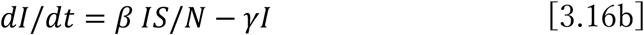

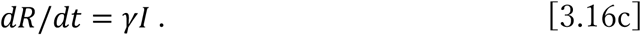

Here *β IS*/*N* is the rate of decrease of the susceptible *S, γ* is the recovery rate (not to be confused with the symbol of a CoV-2 variant strain) and *S* + *I* + *R* = *N*. We assume *N* is a constant (total population of a body we want to discuss). On the other hand, we have Eq. [3.4] which defines the infection numbers in terms of two types of variants *x* and *y* defined by Eqs. [3.1] and [3.2]. By regarding Eqs. [3.16a] and [3.16c] as defining *S* and *R*, the only useful equation is Eq. [3.16b]:

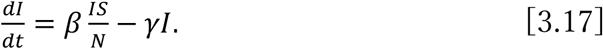

Assuming that the most of the members of the group are susceptible, that is, *S*/*N* ≈ 1, we get

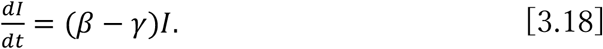

Our idea is that we additively combine epidemiological Eq. [3.18] with genetical Eq. [3.4]. Unlike Bedford et al. (2021), an implicit assumption is that the infection rate is solely determined by pathogen variants and independent of both the host immune history and possible time-dependent antigenic mutation rates. Then we have the equation

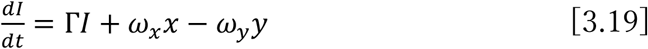

with

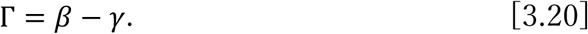

The daily infection number is defined as

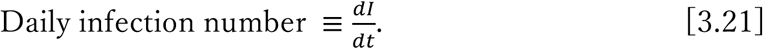

We solve linear differential Eq. [3.19] for *I* using the ordinary method of variation of parameters (in this case it is *f*(*t*)), or we assume *I* in the form of *I*(*t*) = *f*(*t*)Γ*e*^Γ*t*^, substitute it into both sides, and obtain the result of *f*(*t*):

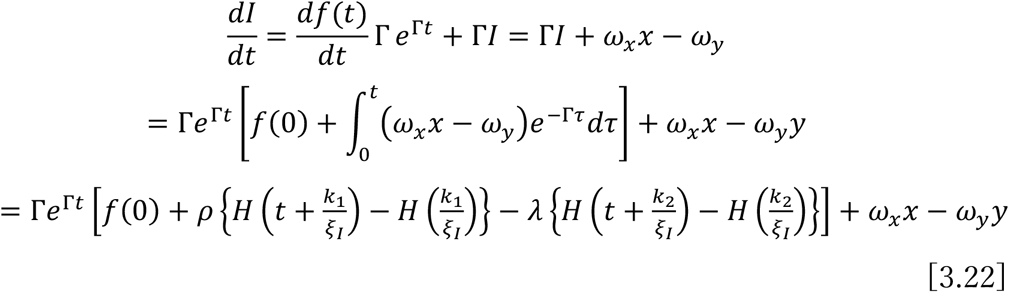

where we used Eq. [3.14] and defined *H*(*x*) as

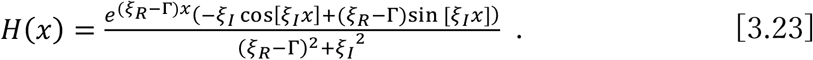

## 4. ANALYSIS OF ACTUAL DATA

In this section, we analyze some data based on Eq. [3.22] with [3.23]. We see that the formula has the generic form:

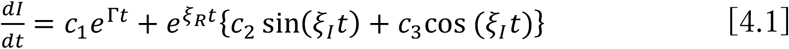

where *c*_*i*_ (*i* = 1, 2, 3) are certain constants. We note that *ξ*_*R*_ and *ξ*_*I*_ are linear functions of *t* as dicussed in the last section. We see that the first term on the right-hand side represents the contribution of the non-genetic factor such as vaccination, body distancing and PCR testing. The second term represents the genetic effect.

### Tokyo 5th wave

We analyze the Tokyo 5th wave data just to see how our new way of analysis works especially in comparison with the standard epidemiology model in Eqs. [3.16]. The Tokyo data on daily infection numbers in the 5th wave period are given in https://www.google.com/search?q=tokyo+data+on+covid-19&oq=tokyo+data&aqs=chrome.1.69i57j35i39j0i512l3j0i30l5.9499j0j15&sourceid = chrome&ie = UTF-8. From the data supplement 1, we have:

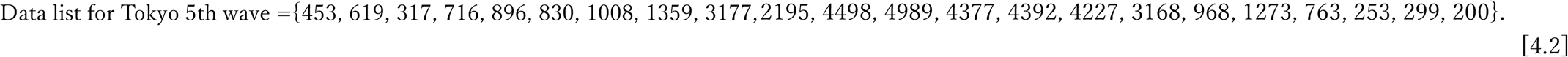

We fit the data with Eq. [4.1] using the nonlinear fitting of Mathematica (Fig. 2). For simplicity, we subsequently denote *ξ*_*R*_ by *R* and *ξ*_*I*_ by *J*. It turned out that we cannot fit the data of the entire period of the 5th wave if we keep the time independent *R*. This is reasonable because the error threshold (*R* = 0) cannot be reached at the start of the spreading. It will be achieved sometime during the spreading process.

**Figure 2.**
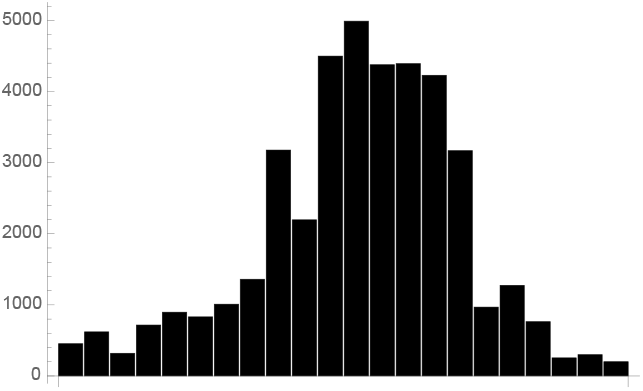
Tokyo daily infection numbers from June 18 to October 1, 2021, (against the horizontal axis *x* measured in units of 5 days, or *x* = *t*/5).

We use two strategies to fit the data:

1. We split the data into three periods and fit the data in each period separately.
2. We assume a simple form of time dependence for *R*. The advantage of this method is that we can calculate from the data when the error threshold is reached.

We start with the method (1): We divide the period of the Tokyo 5th wave into three periods: (a) first 40 days, (b) middle 20 days, and (c) later 40 days. The result of the fitting is as follows:

a. period:

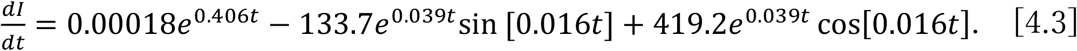 The curve [4.3] is depicted in Fig. 3a. Note that *R* (0.039) is positive indicating that the error threshold has not been reached yet.
b. period: We get

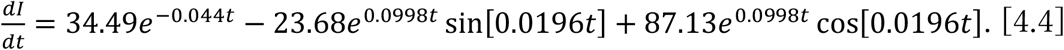 Note that *R* (0.0998) is even larger than the initial period value of 0.039, and it still keeps below the threshold (Fig. 3b).
c. period: In this case we get

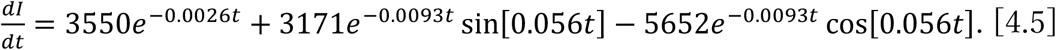

**Figure 3.**
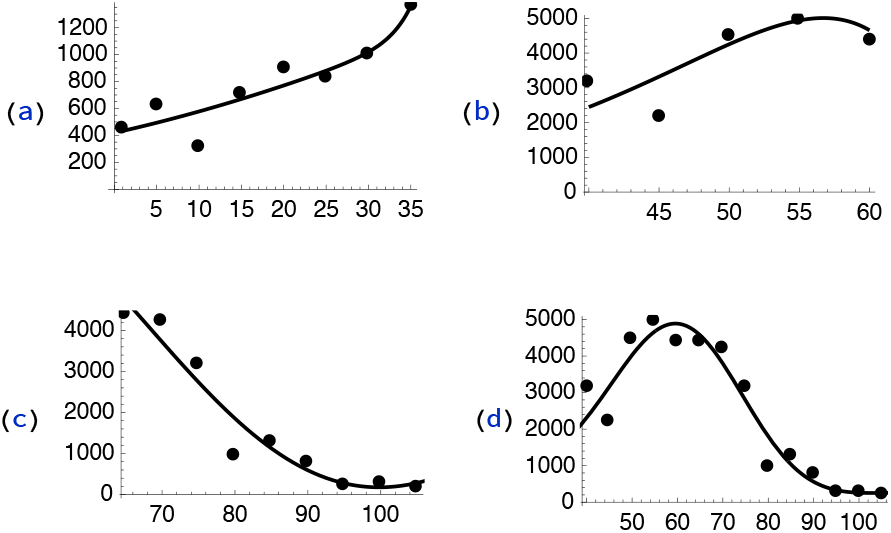
(a) Daily infection number of Tokyo at the initial 40 days of the 5th wave expressed in Eq. [4.3]. (b) Tokyo infection number in the middle 20 days expressed in Eq. [4.4]. (c) Later stage of the Tokyo 5th wave as expressed in Eq. [4.5]. (d) Tokyo 5th wave fit by Eq. [4.9].

We see that *R* (−0.0093) is negative in this period satsfying the error catastrphe condition. Eq. [4.5] is depicted in Fig. 3c.

We learned above that the value of *R* is dependent on which period we consider. This suggests that we may assume some simple form of time dependence for *R* and also for Γ in Eq. [4.2] (see the discussion following Eqs. [3.3] and [3.11a, b] as well as the Appendix concerning symbiosis). The form we assume is the following:

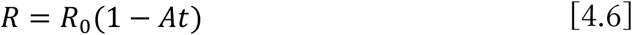

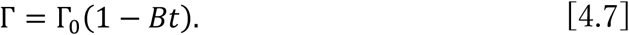

Then, we fit the entire data with a single function:

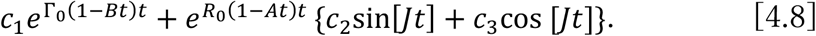

The result of the fitting is given as:

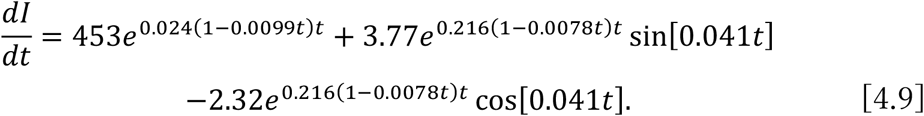

We see from Eq. [4.9] that *R* changes from positive to negative during the 5th wave period. We can calculate the time of going over the error threshold from:

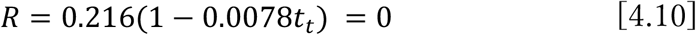

namely, *t*_*t*_ = 128 days after the initiation of the *δ* variant spreading. Although this value of *t*_*t*_ roughly indicates the starting of the decay of the spreading, we see from Eq. [4.9] that other factors and even the non-genetic first term contribute to the precise determination of the maximum position of the infection numbers. Fitting of the entire period by Eq. [4.9] is shown in Fig. 3d. The fitting describes clearly the battle ground of virus vs human immune system. Initially the virus is winning but eventually it reaches the maximum spreading power. The APOBEC3 deaminases involved in the human innate immune system (Robson et al., 2020; Simmonds, 2020; Ratcliff and Simmonds, 2021) keep attacking the virus RNA to keep mutating, which cannot avoid more and more errors so that the spreading power starts to shrink.

## 5. ANALYSIS OF DATA FROM SOUTH AFRICA AND JAPAN

We learned in section 4 that it is possible to fit the data of an entire period of a certain wave if we take into account the time dependence of *R* and Γ (infection rate *β* minus recovery rate *γ* in Eq. [3.20]). We adopt this strategy rather than dividing a period into several sub-periods.

The omicron data are still not sufficient and, therefore, the following analysis should be taken as preliminary rather than final. The omicron variant was first found in South Africa. Therefore, we begin our analysis of the omicron variant in South Africa. The data [https://www.worldometers.info/coronavirus/country/south-africa/] from November 29, 2021 to January 13, 2022 is given as follows (Data supplement 2):

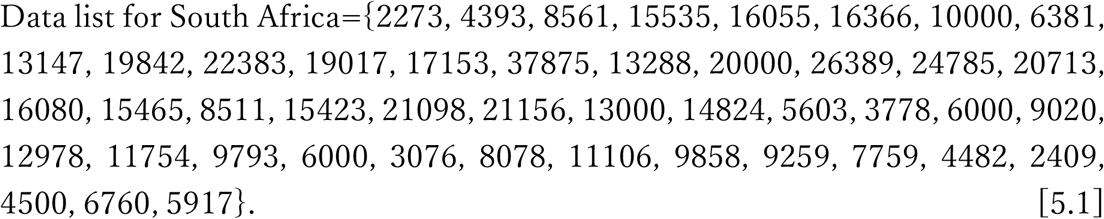

The data is depicted in Fig. 4a. We fit the data with a function that is exactly the same as the one in Eq. [4.8], which is used to explain the entire period of the Tokyo 5th wave. The result of the fitting gives:

**Figure 4.**
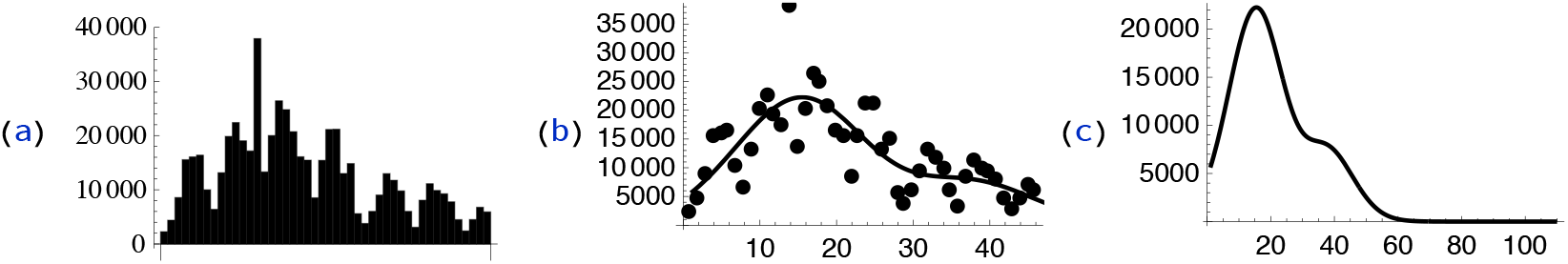
(a) South Africa data of daily infection shown in Eq. [5.1]. (b) South Africa omicron data fit by Eq. [5.2]. (c) Extension of (b) to much later time.

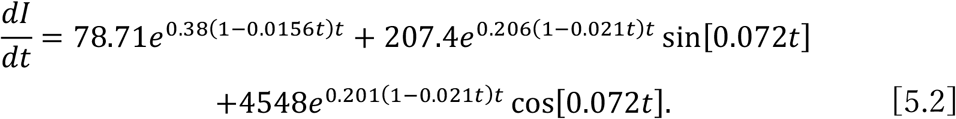

We see from Eq. [5.2] that

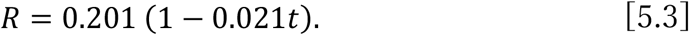

Eq. [5.3] satisfies the error catastrophe condition of *R* < 0 after *t*_*t*_ = 48 days. This value of 48 days is much smaller than the Tokyo 5th wave (supposed to be the *δ* type) value of *t*_*t*_ = 128 days, indicating that omicron infection is more powerful and in turn it reaches the error threshold much faster than the δ type. We show the curve described by Eq. [5.2] in Fig. 4b. We extend Fig. 4b to much later time as in Fig. 4c. We see from Figs. 4b, c that omicron variant infection has a much steeper increase but also sharp decrease due to the earlier arrival at the error threshold.

### Universality of the parameters ξ_***R***_ (= ***R***) and ξ_***I***_ (= ***J***)

Before fitting the Japan data, we discuss the nature of the parameters *ξ*_*R*_ and *ξ*_*I*_. As shown above the time to reach the error threshold is calculated using the time dependence of *ξ*_*R*_. We obtained the value *t*_*t*_ = 128 days for the *δ* type using the Tokyo data and *t*_*t*_ = 48 days for the omicron variant using the South Africa data. We assume that these values depend on the variants we consider, but not or very little on any other external effect such as vaccination, social distancing etc.. The reason is that *ξ*_*R*_ and *ξ*_*I*_ are genetic parameters but not the epidemic parameters. They are more or less inherent in the RNA of the virus and can also depend on the genetically determined human immune system. The other parameters in Eq. [4.1] can depend on vaccination, social distancing, PCR testing followed by segregation etc.. The above argument allows us to use the values of *ξ*_*R*_ and *ξ*_*I*_ obtained from the South Africa data to analyze the omicron data of other regions, although there may be some difference of values of *ξ*_*R*_ and *ξ*_*I*_ due to population structure such as how much of particular genomic segments of Neanderthals introgressing into the DNA of the group we deal with (Zeberg and Pääbo, 2020). In the following analysis of the Japan data, we use the South Africa *ξ*_*R*_ and *ξ*_*I*_ for simplicity.

The validity or the usefulness of the universality of *ξ*_*R*_ and *ξ*_*I*_ can be checked by the analysis using such index as the AIC. Along the same line, we can also consider (calculate the AIC of) the limiting case of all the genetic part of the parameters vanishing. These works are left to our future publication.

### Japan’s case -- 6th wave

We analyze the Japan data of daily infection number starting December 30, 2021 to January 13, 2022 when the omicron variant was starting to dominate. We use the same *ξ*_*R*_ and *ξ*_*I*_ obtained in the South Africa analysis. The data for the 6th wave of Japan is given in the data supplement 3 as,

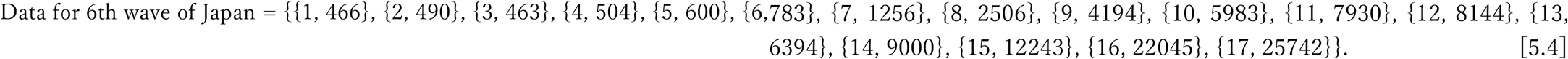

We fit this with the following function which is exactly same as the one used in the South Africa analysis with an unjustified assumption that even the non-genetic parameter (*B* in Eq. [4.7]) is universal:

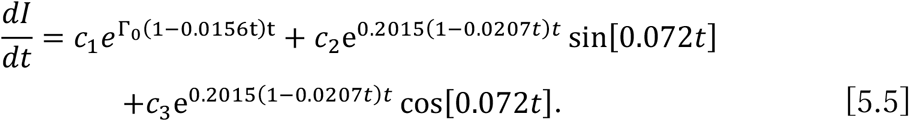

The result of the fitting gives

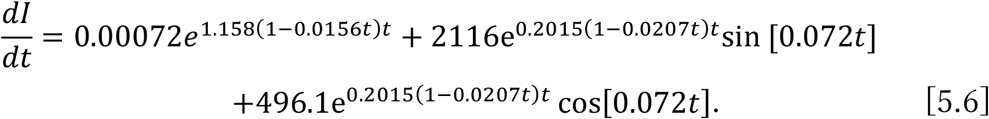

The data [5.4] and Eq. [5.6] are plotted in Fig. 5a. From Fig. 5b, we see that Japan’s daily infection number will go up to approximately 100,000 and then it starts decreasing. Our analysis of the Japan data (dataJPN1 in [5.4]) from December 30, 2021 to January 13, 2022 correctly predicted the maximum daily infection number to be approximately 100,000. But the actual speed of the decline from the peak number is much less than predicted by the analysis. This is due to the unjustified assumption that even the non-genetic parameters are universal.

**Figure 5.**
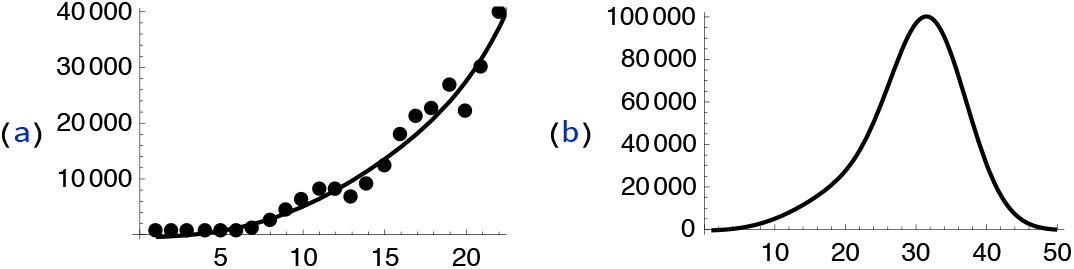
(a) Japanese daily infection number fitted by Eq. [5.6]. (b) Extension of (a) to later time.

### Analysis of later data of Japan

We investigate the situation by analyzing the data from January to March, 2022. Here we drop the unjustified assumption that the non-genetic parameters are universal. The data from January 1 to March 31 of the infection number (every 3 days) is given in the data supplement 4 as,

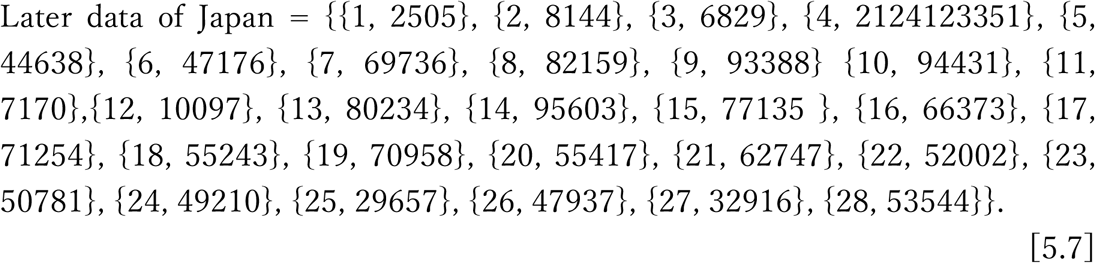

We fit the data with the following function:

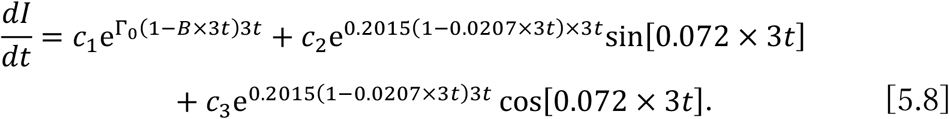

The meaning of the right-hand side in Eq. [5.8] is the following:

1. Compared to the previous fitting curve given in Eq. [5.5], we see that we use “3*t*” rather than “*t*” to designate the time variable (here we have data points of every 3 days).
2. Otherwise, the only difference is the insertion of the parameter *B* in the first term (social distancing contribution). This is the most important deviation from Eq. [5.5] where the unjustified assumption of the non-genetic term was made.
3. The universality of the mutation contribution is maintained: The second and third terms are the same as in Eq. [5.5].

The result of the fitting gives:

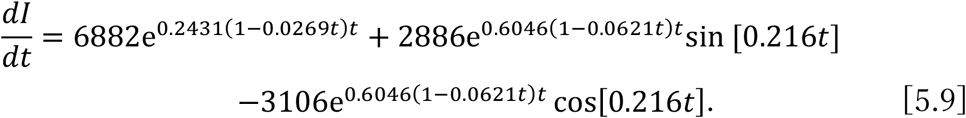

This non-linear fitting is shown in Fig. 6a. We extend the function in Eq. [4.9] to *t* = 50 (150 days after January 1, 2022, the end of May). It is depicted in Fig. 6b.

**Figure 6.**
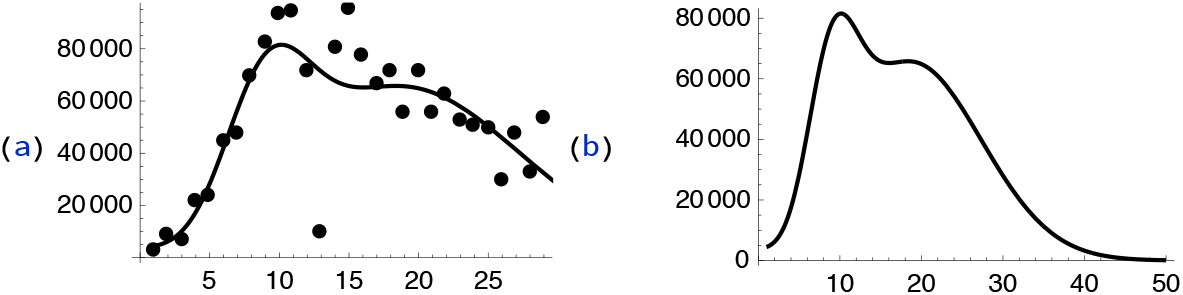
(a) Fitting of the Japan data of daily infection from January 1 to April 2 fitted by Eq. [5.9]. (b) Daily infection number of Japan from early January to the end of May given in Eq. [5.9].

### Tentative summary of the analysis of the later data of Japan

(1) As one can see by comparing Eqs. [5.6] and [5.9], the difference between the previous analysis and the present one is mostly in the first term representing the contribution of the social distancing effect. The previous best fitting ended up in minimizing the first term, whereas the current fitting has much larger contribution from this term.

(2) The exponential factor of the first term in the current analysis 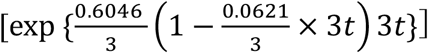 behaves more or less the same way as that of the previous analysis [exp {0.2015(1 − 0.0207*t*)*t*}] indicating the structure of the social distancing effect was correctly taken into account in both analyses. The difference is in the overall factor of the first term (0.00072 vs 6882).

(3) As one can see from Fig. 6b, there will be no 7th wave in Japan if our analyses are correct. Unfortunately, however, it appears that before omicron BA. 2 completely disappears, it was gradually replaced by a new omicron variant strain (BA. 5), which has caused the 7th wave in 2022 summer Japan.

One of the authors (H. S.) would like to express his sincere gratitude to Professor Toshimichi Ikemura for his variable discussions and guidance.

## Data Availability

All data produced in the present work are contained in the manuscript and available online at 
https://www.google.com/search?q=tokyo+data+on+covid-
19&oq=tokyo+data&aqs=chrome.1.69i57j35i39j0i512l3j0i30l5.9499j0j15&sourceid = chrome&ie = UTF-8,
https://www.worldometers.info/coronavirus/country/south-africa/,
https://www.worldometers.info/coronavirus//country/japan/.

https://www.google.com/search?q=tokyo+data+on+covid-19&oq=tokyo+data&aqs=chrome.1.69i57j35i39j0i512l3j0i30l5.9499j0j15&sourceid=chrome&ie=UTF-8

https://www.worldometers.info/coronavirus/country/south-africa/

https://www.worldometers.info/coronavirus//country/japan/

## APPENDIX SYMBIOSIS OR TOTAL EXTINCTION

Our definition of error catastrophe discussed in section 2 is given in terms of the real part of the eigenvalue of the matrix which determines the numbers of variants which contribute positively or negatively to the infection of a given virus. The eigenvalue is time dependent in general. If the real part is positive, the positive variant type dominates and vice versa. The error catastrophe occurs when the real part becomes zero at certain stage. If the negativity persists, we will reach the total extinction of the virus.

Now suppose that the real part of the eigenvalue stays zero after reaching the error catastrophe. This is the situation of the symbiosis due to our definition. Then our basic Eq. [4.1] becomes,

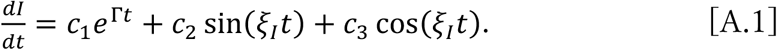

The first term is the contribution from the artificial measure such as vaccination, social distancing etc.. The rest shows the mutation effect. The value of Γ can be negative without any of the artificial measures corresponding to the weakened infection power of the virus. It is possible that we still need vaccination etc. to make it negative.

On the mutation effect, we see that the daily infection number oscillates as a function of time if the imaginary part of *ξ*_*I*_ (= *J*) is non-zero. This implies that, in case of symbiosis, there will be maximum and minimum in the infection rate somewhat similar to the situation where the infection rate reached the error catastrophe. But the difference is that the former is realized due to the non-zero property of *ξ*_*I*_, whereas the latter is due to the vanishing of *ξ*_*R*_ (= *R*).

This means that there could be ups and downs of daily infection numbers in the case of symbiosis, as in the case of seasonal influenza, and this phenomenon is not the same as in the case of error catastrophe.

### Data Supplement

1. Data list for Tokyo 5th wave Our format of the data is such that we list the number of patients starting from June 18 to October 1, 2021 every 5 days: Data list for Tokyo 5th wave ={453, 619, 317, 716, 896, 830, 1008, 1359, 3177, 2195, 4498, 4989, 4377, 4392, 4227, 3168, 968, 1273, 763, 253, 299, 200}.
2. Data list for South Africa Our format of the data here is such that we list the number of patients starting from November 29, 2021 to January 13, 2022 every day: Data list for South Africa={2273, 4393, 8561, 15535, 16055, 16366, 10000, 6381, 13147, 19842, 22383, 19017, 17153, 37875, 13288, 20000, 26389, 24785, 20713, 16080, 15465, 8511, 15423, 21098, 21156, 13000, 14824, 5603, 3778, 6000, 9020, 12978, 11754, 9793, 6000, 3076, 8078, 11106, 9858, 9259, 7759, 4482, 2409, 4500, 6760, 5917}.
3. Data for 6th wave of Japan Here the same data format as in the case of South Africa is used except that we add the numbering of dates on the first column of the list: Data for 6th wave of Japan = {{1, 466}, {2, 490}, {3, 463}, {4, 504}, {5, 600}, {6, 783}, {7, 1256}, {8, 2506}, {9, 4194}, {10, 5983}, {11, 7930}, {12, 8144}, {13, 6394}, {14, 9000}, {15, 12243}, {16, 22045}, {17, 25742}}.
4. The Japan data from January 1 to March 31 of the infection number (every 3 days) (https://www.worldometers.info/coronavirus//country/japan/) (using the same format as in the case of 6th wave): Later data of Japan = {{1, 2505}, {2, 8144}, {3, 6829}, {4, 2124123351}, {5, 44638}, {6, 47176}, {7, 69736}, {8, 82159}, {9, 93388} {10, 94431}, {11, 7170},{12, 10097}, {13, 80234}, {14, 95603}, {15, 77135}, {16, 66373}, {17, 71254}, {18, 55243}, {19, 70958}, {20, 55417}, {21, 62747}, {22, 52002}, {23, 50781}, {24, 49210}, {25, 29657}, {26, 47937}, {27, 32916}, {28, 53544}}.

## Notes

### Competing Interest Statement

The authors have declared no competing interest.

### Funding Statement

This study did not receive any funding

### Author Declarations

The study used ONLY oepnely available human data that were originally located at: https://www.google.com/search?q=tokyo+data+on+covid- 19&oq=tokyo+data&aqs=chrome.1.69i57j35i39j0i512l3j0i30l5.9499j0j15&sourceid = chrome&ie = UTF-8, https://www.worldometers.info/coronavirus/country/south-africa/, https://www.worldometers.info/coronavirus//country/japan/.

### Summary of Updates

We clarified that our main aim of this paper is to model genetic error catastrophe in a simple manner. For this purpose, we briefly surveyed current studies of epidemiology with special reference to roles of antigenic mutations and emergence of new variant strains. Although our model formalized as in Eq. [3.19] does not assume any interaction between host and pathogens, we emphasized that our formalism can be regarded as a starting point for integrating genetics into epidemiology. Accordingly we revised the abstract, introduction and conclusion. One such is the following phrases inserted in the introduction: A simplified assumption of the absence of mutations in the virus genome is still made even for recently developed models in which, for example, multiple pathogen strains interact to each other through cross-immunity (Kucharski, Andreasen and Gog, 2016 for a review) or strains may interfere through the removal of individuals from the susceptible pool after an acute infection (e.g., Rohani et al., 2002). Meanwhile, the importance of random nature of pathogen mutations and emergence of new strains has been well recognized and studied by simulation under the assumption that a host risk of infection is proportional to the genetic distance between the infecting phenotype and the closest phenotype in the host immune history (Bedford et al., 2012). Recently, Miller, Elenberg and Dubrawski (2022) explored an existing agent-based model for influenza A/H3N2 which incorporates mutation effects into both transmission rate and the antigenic space in the standard model. We present here a way to remedy the current situation for the presence of largely deleterious but occasionally advantageous antigenic mutations by introducing into the scheme of the standard model some elements showing the vital role played by mutations before, during and after the period of extensive virus spreading.

## REFERENCES

Bedford, T., Rambaut, A., and Pascual, M. (2012) Canalization of the evolutionary trajectory of the human influenza virus. BMC biology 10: 38.

Eigen, M., (1971) Self-organization of matter and the evolution of biological macro-molecules. Naturwissenschaften 58: 465–523.

Kermack, W. O., and McKendrick, A. G. (1927) A contribution to the mathematical theory of epidemics. Proc. R. Soc. Lond. A 115: 700–721.

Kucharski, A. J., Andreasen, V., and Gog, J. R. (2016) Capturing the dynamics of pathogens with many strains. J. Math. Biol. 72: 1–24.

Miller, J. K., Elenberg, K., and Dubrawski, A. (2022) Forecasting emergence of COVID-19 variants of concern. PLoS ONE 17(2): e0264198.

Ratliff, J., and Simmonds, P. (2021) Potential APOBEC-mediated RNA editing of the genomes of SARS-CoV-2 and other coronaviruses and its impact on their longer term evolution. Virology 556: 62–72.

Robson F., Khan, K. S., Paris, C., Demirbag, S., Barfuss, P., Rocchi, P., and Ng, W. L. (2020) Coronavirus RNA proofreading: Molecular basis and therapeutic targeting. Mol. Cell 80: 1136–1138.

Rohani, P., Green, C. J., Mantilla-Beniers, N. B., and Grenfell, B. T. (2002) Ecological interference between fatal diseases. Nature 422: 885–888.

Sadeghpour, S., Khodaee, S., Rahnama, M., Rahimi, H., and Ebrahimi, D. (2021) Human APOBEC3 variations and viral Infection. Viruses 13: 1366. https://doi.org/10.3390/v13071366.

Simmonds, P. (2020) Rampant C→U hypermutation in the genomes of SARS-CoV-2 and other coronaviruses: Causes and consequences for their short- and long-term evolutionary trajectories. mSphere 5: e00408–20.

Smith, E. C., Blanc, H., Vignuzzi, M., and Denison, M. R. (2013) Coronaviruses lacking exoribonuclease activity are susceptible to lethal mutagenesis: Evidence for proofreading and potential therapeutics. PLoS Pathog 9(8): e1003565. doi: 10.1371/journal.ppat.1003565.

Sugawara, H. (2021a) On the effectiveness of the search and find method to suppress spread of SARS-CoV-2. Proc. Jpn. Acad. Ser. B 97(1): 22–49.

Sugawara, H. (2021b) Effect of vaccination against COVID-19 spreading. Proc. Jpn. Acad. Ser. 97(9): 543–558.

Thakur, V., Bhola, S., Thakur, P., Patel, S. K. S., Kulshrestha, S., Ratho, R. K., and Kumar, P. (2022) Waves and variants of SARS-CoV-2: understanding the causes and effect of the COVID-19 catastrophe. Infection 50: 309–325.

Zeberg, H., and S. Pääbo (2020) The major genetic risk factor for severe COVID-19 is inherited from Neanderthals. Nature 587: 610–612.

